# Esophageal Cooling For Protection During Left Atrial Ablation: A Systematic Review And Meta- Analysis

**DOI:** 10.1101/19003228

**Authors:** Lisa Leung, Mark Gallagher, Pasquale Santangeli, Cory Tschabrunn, Jose Guerra, Bieito Campos, Jamal Hayat, Folefac Atem, Steven Mickelsen, Erik Kulstad

## Abstract

**Background:** Thermal damage to the esophagus is a risk from radiofrequency (RF) ablation of the left atrium for the treatment of atrial fibrillation (AF), with the most extreme type of thermal injury resulting in atrio-esophageal fistula (AEF), with a correspondingly high mortality rate. Various approaches have been developed to reduce esophageal injury, including power reduction, avoidance of greater contact-force, esophageal deviation, and esophageal cooling. One method of esophageal cooling involves direct instillation of cold water or saline into the esophagus during RF ablation. Although this method provides limited heat-extraction capacity, studies of it have suggested potential benefit.

**Objective:** We sought to perform a meta-analysis of existing studies evaluating esophageal cooling via direct liquid instillation for the reduction of thermal injury.

**Methods:** We reviewed Medline for existing studies involving esophageal cooling for protection of thermal injury during RF ablation. A meta-analysis was then performed using random effects model to calculate estimated effect size with 95% confidence intervals, with outcome of esophageal lesions, stratified by severity, as determined by post-procedure endoscopy.

**Results:** A total of 9 studies were identified and reviewed. After excluding pre-clinical and mathematical model studies, 3 were included in the meta-analysis, totaling 494 patients. Esophageal cooling showed a tendency to shift lesion severity downward, such that total lesions did not show a statistically significant change (OR 0.6, 95% CI 0.15 to 2.38). For high grade lesions, a significant OR of 0.39 (95% CI 0.17 to 0.89) in favor of esophageal cooling was found, suggesting that esophageal cooling, even utilizing a low-capacity thermal extraction technique, reduces lesion severity from RF ablation.

**Conclusions:** Esophageal cooling reduces lesion severity encountered during RF ablation, even when using relatively low heat extraction methods such as direct instillation of cold liquid. Further investigation of this approach is warranted.

## INTRODUCTION

Thermal damage to the esophagus is a risk from radiofrequency (RF) ablation or cryoablation of the left atrium for the treatment of atrial fibrillation (AF).[1-3] The most extreme type of thermal injury is an atrio-esophageal fistula (AEF), with a mortality rate of 80% or more.[4-8] Various approaches have been developed to reduce esophageal injury, including power reduction, avoidance of greater contact-force, temperature monitoring, esophageal deviation, and esophageal cooling. Different esophageal protection approaches have been evaluated including temperature monitoring, esophageal deviation, and power reduction, with varying degrees of success in reducing esophageal injury.[9-11]

Esophageal cooling has also been investigated in multiple studies for the purpose of protecting the esophagus during RF ablation. [12-20] Various approaches to cooling have been utilized, including expandable balloon devices, a cooling sac circulating water, and direct instillation of ice-cold water or saline into the esophagus, with investigations utilizing animal models, mathematical models, and human clinical studies. The majority of human clinical studies have utilized direct instillation of ice cold water or saline. We sought to perform a meta-analysis of these existing data to examine the range of effect sizes published to date and estimate the potential efficacy of esophageal cooling for protection during RF ablation.

## Methods

### Data sources and search strategy

We utilized PubMed to perform a search of the literature from 1985 (prior to the earliest reports of endocardial ablation to treat atrial fibrillation) to June 2019 for studies published on esophageal cooling during cardiac ablation using a broad search with the following Boolean structure: (esophag* OR oesophag*) AND cooling AND (ablation OR fibrillation) We did not restrict to English only. Details of the systematic review were submitted for registration in PROSPERO on June 21, 2019.

### Eligibility criteria

We excluded pre-clinical studies, bench-top, agar phantom, and mathematical model studies, and studies that did not include formal endoscopy as an outcome measure.

### Data collection

The primary data of interest were endoscopically identified lesions found after RF ablation. Because we anticipated inconsistency in the categorization of lesion severity, we decided to simplify all lesion severity measurement into severe lesions characterized by the presence of ulceration and mild to moderate lesions encompassing all other abnormalities.

### Statistical analysis

We input study data into Review Manager 5.3 to perform meta-analysis of the data entered, and present the results graphically. SAS version 9.4 (SAS Institute Inc., Cary, NC, USA) was used for additional analyses. The Cochran-Mantel-Haenszel (CMH) method was employed to test the null hypothesis that the response rate is the same for the two arms (control versus treatment), after adjusting for possible differences in study response rates. Furthermore, we fitted a random effect model using SAS procedures GLIMMIX and NLMIXED by treating the study as a random effect. Because lesion grades are often considered to be dichotomized into those that are likely to progress to atrio-esophageal fistula, and those that are not, we initially analyzed the data as a binary outcome (high grade lesions concerning for progression versus low grade lesions likely to heal spontaneously). Then, to further estimate effect size, we used an ordinal logistic random intercept model, taking into account the ordered nature of lesion grading (low to high, numerically).

## RESULTS

A total of 9 studies were identified using the above criteria and additional search strategy. Five of these were excluded for being non-clinical. Berjano et al. utilized a finite element model in three dimensions to investigate the effects of a cooled intra-esophageal balloon.[12] Lequerica et al. performed studies using an agar phantom-based model that was built to provide temperature readings at points between the esophageal lumen and the myocardium.[13, 14] Arruda et al. studied a custom developed system utilizing temperature controlled saline or water in an in vitro lamb heart and esophagus preparation, followed by an in vivo model with six dogs.[15] Scanavacca et al. presented a study of the use of a saline filled esophageal balloon to attempt esophageal protection in a dog model.[17] A clinical study by Tsuchiya et al. involving 8 patients was excluded for not utilizing endoscopy to determine the presence of lesions after RF ablation.[16]

The remaining 3 studies included a total of 494 patients:

John et. al performed a study of 76 patients, half given active cooling performed by injecting a 20 mL bolus of ice-cold saline via orogastric tube into the upper esophagus if/when the luminal esophageal temperature increased by 0.5 °C above baseline.[20] The authors found that this method of esophageal cooling did not decrease the overall incidence of thermal lesions, but noted a trend toward fewer severe lesions with cooling (Fig 1). Esophageal thermal lesions were graded as follows: grade 0: no esophageal lesion; grade 1: mucosal damage <1 cm width; grade 2: mucosal damage 1–3 cm width; grade 3: mucosal damage > 3 cm width or visualization of deeper layer; and grade 4: bleeding ulcer or with overlying clot.

**Figure 1.**
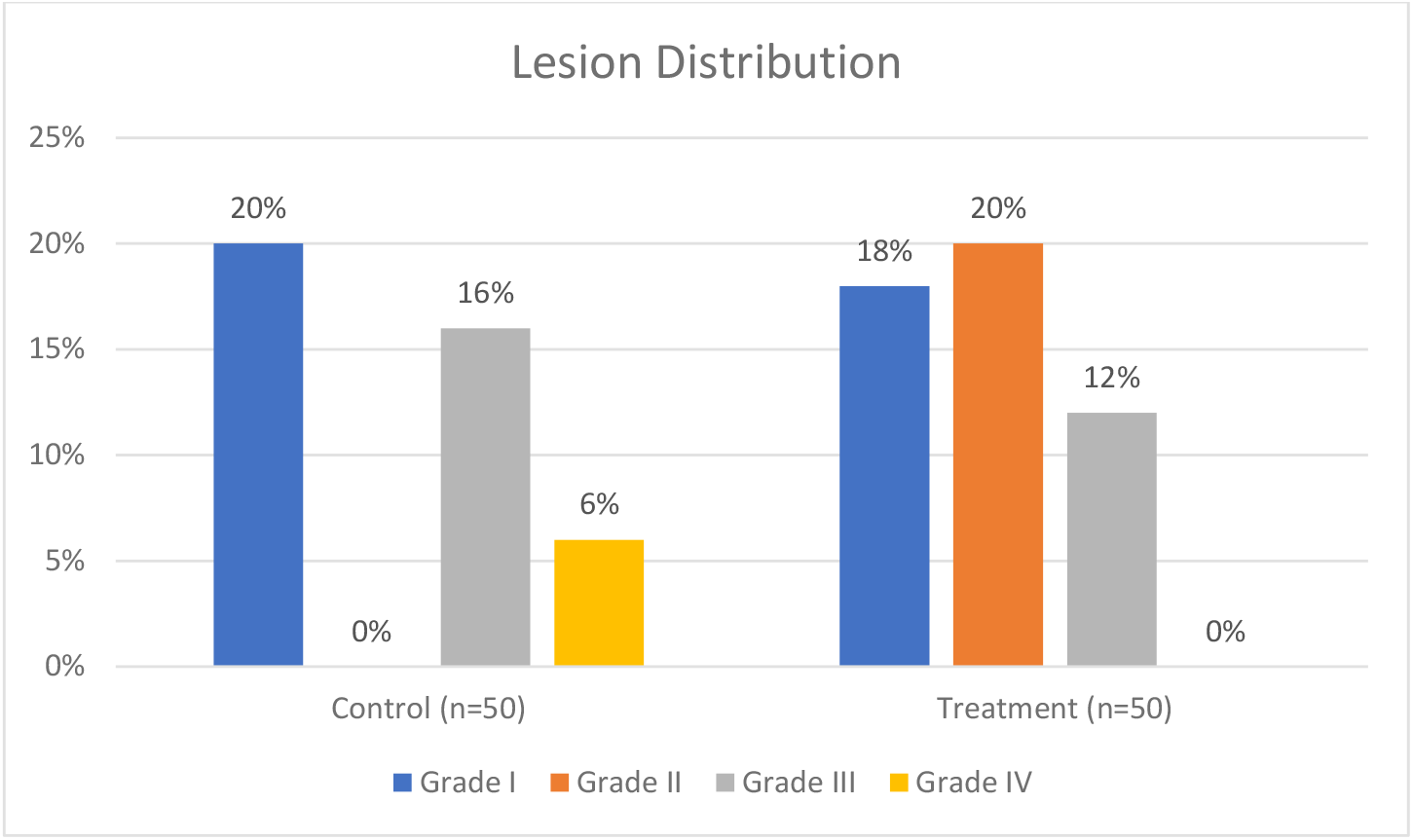
Results from John et al. Grade III and grade IV lesions are shown separately.

Kuwahara et. al performed a study of 100 patients using very small volumes (5 mL) of ice water as the which was injected prior to RF energy delivery, and subsequently when esophageal temperatures reached 42°C.[18] The severity of the esophageal lesions was qualitatively graded as mild, moderate, or severe, according to their extent and color, with the severe category corresponding to those categorized as grade III or IV by John et al. The authors found that this approach reduced the severity of esophageal lesions, but did not reduce their incidence: lesions occurred in 20% of the experimental group, and 22% of the controls, with 3 moderate and 7 mild in the cooled group and 3 severe, 1 moderate, and 7 mild in the control (Fig 2).

**Figure 2.**
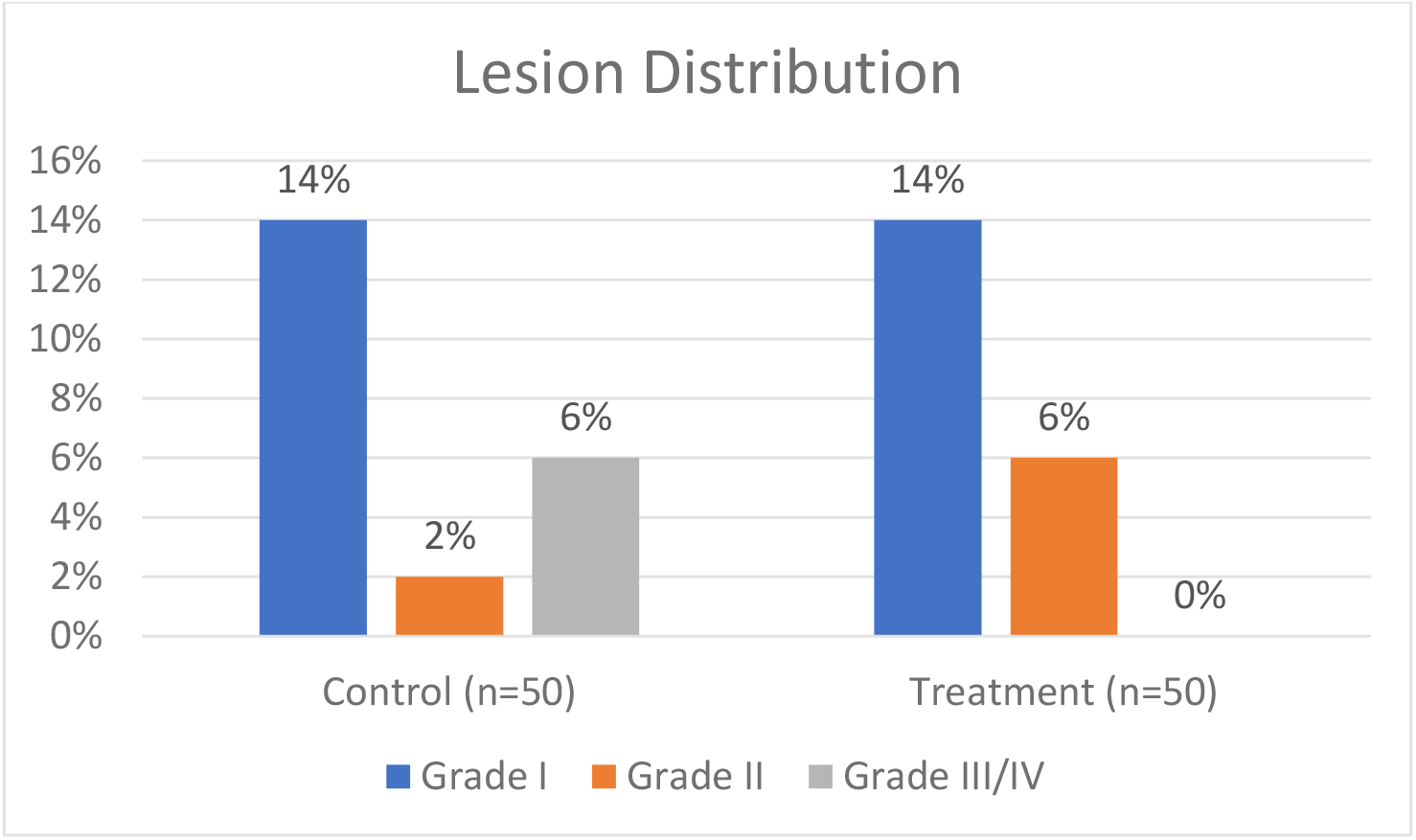
Results of Kuwahara et. Al. The Grade III/IV lesion category represents all those qualitatively graded as “severe”, with mild lesions in Grade I and moderate lesions in Grade II.

Sohara et. al utilized an infusion of cooled saline mixed with Gastrografin or Iopamidol, with slightly higher, but still limited volumes (10 – 20 mL) in repeated injected aliquots with a temperature of approximately 10°C.[19] A total of 318 patients were randomized between groups receiving only temperature monitoring without cooling, temperature monitoring with cooling when temperature exceeded 43°C, and temperature monitoring with cooling received when temperatures exceeded 39°C. The percentage of patients free from any ulceration or erosion in each group was found to be 63.6%, 87.5%, and 95.2%, respectively (Fig 3). Esophageal lesions were classified as normal (score 1), erosion (patchy mucosal ulceration: score 2), mild ulcer (necrosis less than 3 mm in diameter with red spot: score 3), severe ulcer (necrosis more than 3 mm in diameter with red spot and/or with a hemorrhagic appearance, often with fibrinoid material: score 4).

**Figure 3.**
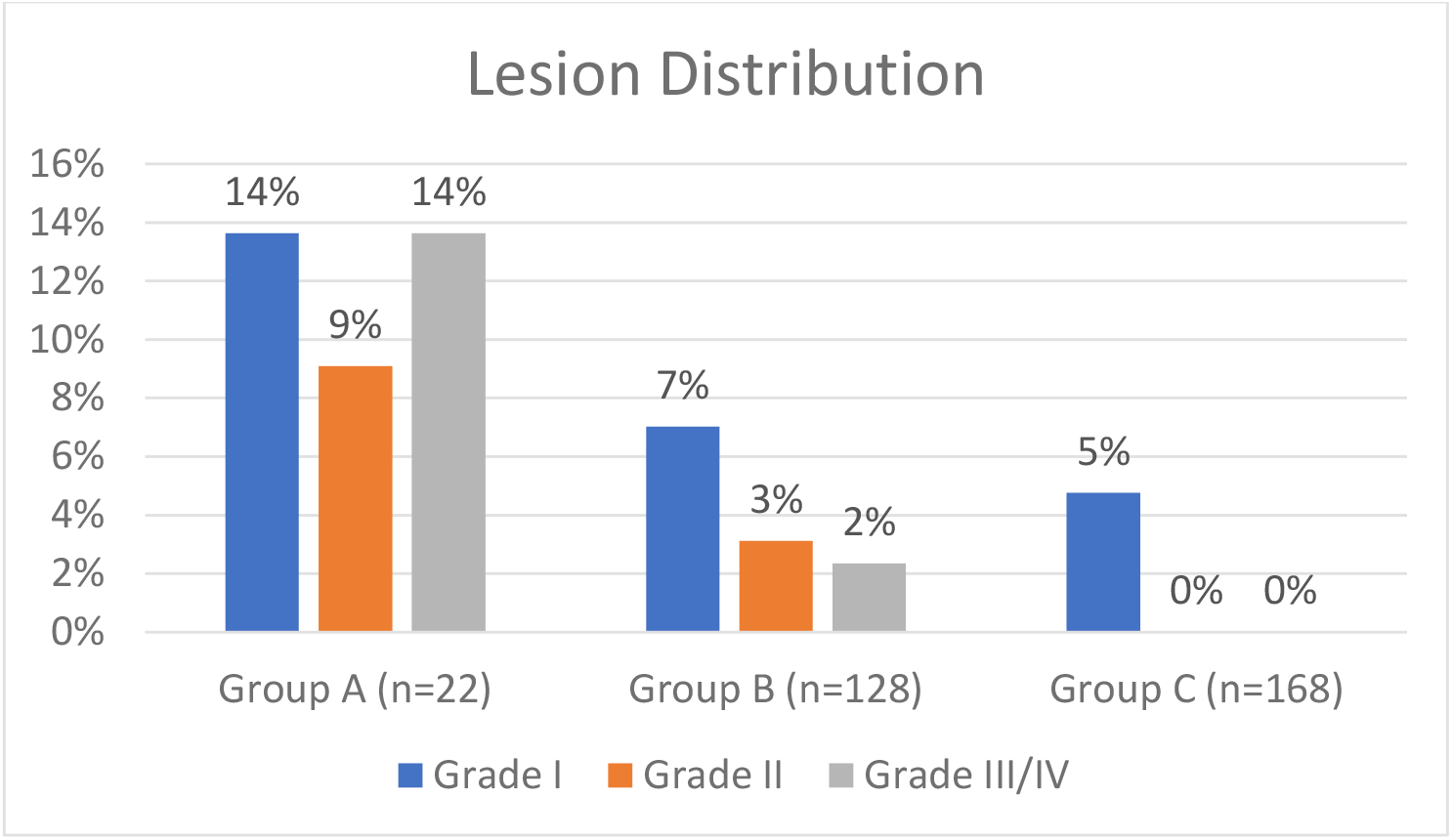
Results from Sohara et al in their study of 318 consecutive patients. Group A had local temperature monitoring without cooling of the esophagus. Groups B and C patients had monitoring with cooling of the esophagus when the LET exceeded 43 °C and 39 °C, respectively. The Grade III/IV lesion category represents all those graded as ulcers (score 3 or 4 in the author’s categorization system).

In 2 of these 3 studies, a clear shift from high-grade to lower-grade lesions can be seen between the control and treatment arms.[18, 20] In contrast, in Sohara et al., a general reduction of all grade lesions is seen.[19]Further details on the characteristics of each study are shown in Tables 1, 2, and 3.

**Table 1.**
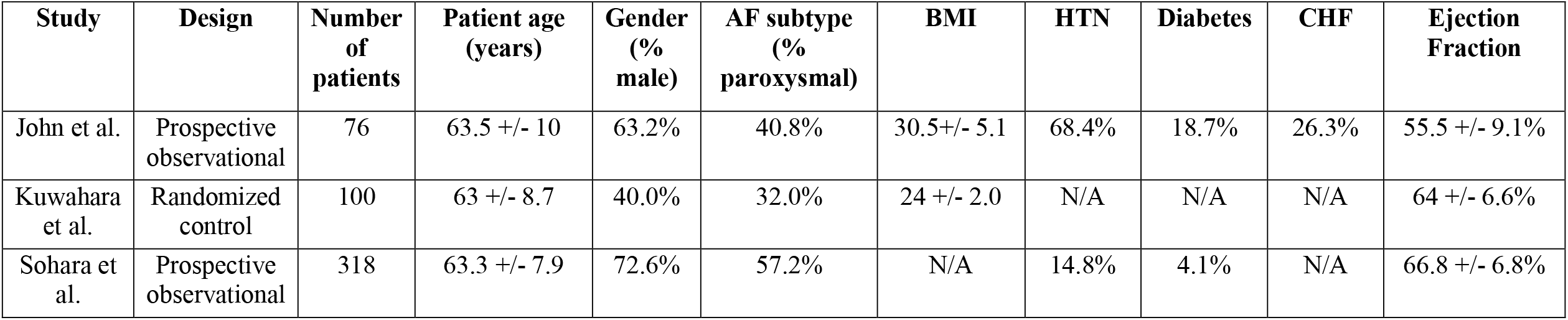
Patient characteristics and study type. Group data from each study averaged over all groups.

**Table 2.** Ablation characteristics and technology.

| Study | Ablation technology | Power | Mapping technology | Ablation type | Average Contact Force | Mean RF time per lesion (seconds) | Anesthesia | Endoscopy timing |
| --- | --- | --- | --- | --- | --- | --- | --- | --- |
| John et al. | RF open-irrigation, 3.5-mm-tip, 8-Fr, force-sensing catheter (SmartTouch) | 24-28 W | CARTO mapping system and PentaRay NAV multipolar mapping catheter | Bilateral antral PVI, creation of a left atrial roof and floor line, and ablation across the mitral isthmus | 10 g | 12.3 +/- 5.3 | General | Within 24 hours of the procedure |
| Kuwahara et al. | 3.5 mm irrigated-tip ablation catheter (Thermocool, Biosense Webster, Inc.) | 25-30 W | CARTO system | Circumferential PVI with focal ablation, and creation of left atrial roof or mitral isthmus lines with posterior wall isolation in non-paroxysmal AF | N/A | N/A | Conscious sedation | Within 24 hours of the procedure |
| Sohara et al. | 12F radiofrequency hot balloon catheter (Hayama Arrhythmia Institute, Kanagawa, Japan) | N/A | CARTO system | Balloon-based box isolation | N/A | N/A | General | Within 3 days of the procedure |

**Table 3.** Esophageal cooling technique.

| **Study** | **Temperature sensor type** | **Temperature sensor characteristics** | **Gastric tube type** | **Coolant** | **Cooling threshold** | **Follow up duration** |
| --- | --- | --- | --- | --- | --- | --- |
| John et al. | 18-Fr esophageal temperature probe (400 series M1024215, GE Healthcare, Chicago, IL) | Tube diameter 6 mm, cuff diameter 8.8 mm, single thermistor inside of the cuff at the distal tip | 18-Fr orogastric tube (Nasogastric Sump Tube 0046180, Bard, Inc., Covington, GA) | 20mL ice-cold saline | 0.5°C increase in temperature from baseline | N/A |
| Kuwahara et al. | A multi-thermocouple temperature probe (Sensitherm, St Jude Medical) | Three thermocouples; one of which was placed at the same level of the esophagus as the site of RF energy delivery on the LA posterior wall | Unspecified gastric tube | 5mL ice-water (0 °C) | 42°C temperature peak | Up to 8 weeks |
| Sohara et al. | Thermocouple thermometer (Delta Ohm, Caselle di Selvazzano, PD, Italy) | Thermal sensor of a deflectable, 4-mm tip ablation catheter | Unspecified gastric tube coated with xylocaine jelly | 10-20mL ionized contrast medium (Gastrografin) or nonionized low osmotic contrast medium (iopamidol) diluted 1:2 with physiologic saline refrigerated to about 10 °C | 43°C temperature peak (Group B) or 39°C temperature peak (Group C) | >10 months minimum, mean 3.6 years |

The forest plot comparing the outcome of all lesions (grades I, II, III, and IV) as events between control and treatment arms is shown in Fig 4. Although fewer lesions occurred in the treatment arms of 2 of the 3 studies, in meta-analysis this did not reach statistical significance (OR 0.6, 95% CI 0.15 to 2.38).

**Figure 4.**
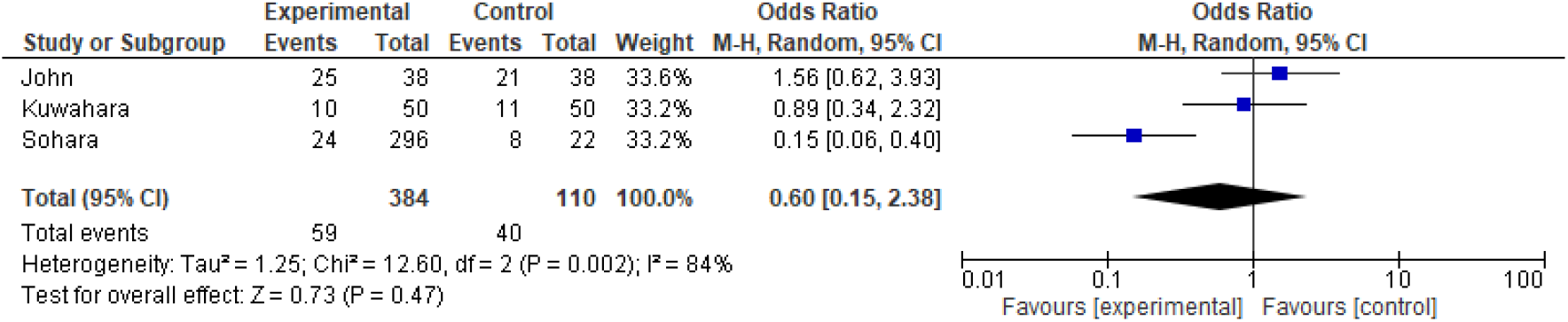
Forest plot of clinical studies comparing outcome of all lesions. Events are the occurrence of grade I, II, III, and IV lesions.

Further depiction of the slight increase in low grade lesions seen in 2 of the 3 studies is shown in the forest plot in Figure 5, with an OR of 1.0 (95% CI 0.26 to 3.93) suggesting that overall, low grade lesions are not significantly impacted with this treatment.

**Figure 5.**
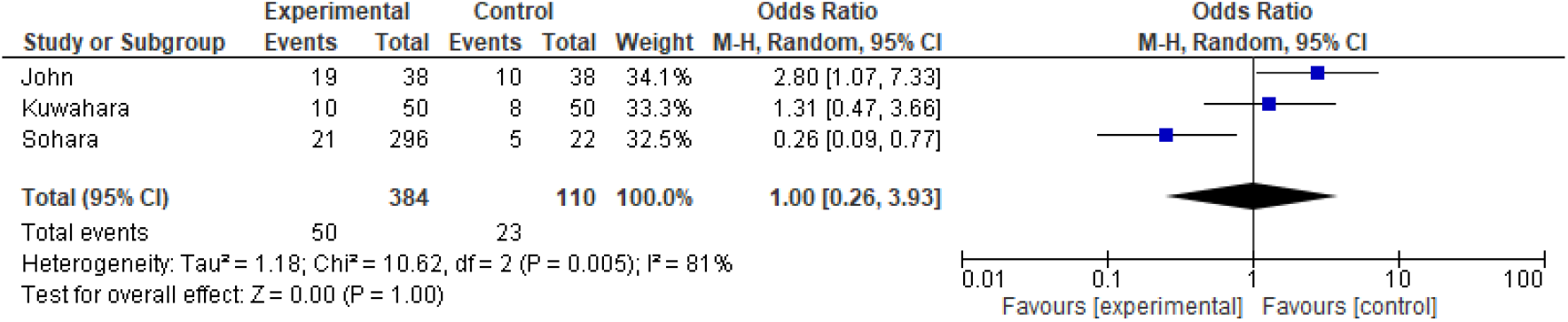
Forest plot of clinical studies comparing outcome of low-grade lesions. Events are the occurrence of grade I and II lesions.

Evaluating the occurrence of high-grade lesions (grade III and IV) results in the forest plot shown in Fig 6, demonstrating a significant OR of 0.39 (95% CI 0.17 to 0.89) in favor of the treatment arm. Separately, utilizing the CMH method, we obtained a significant p-value of 0.016 indicating the association between treatment and grade remains strong. Furthermore, in a binary logistic regression model, an OR of 0.46 (95% CI 0.28 to 0.75) was found.

**Figure 6.**
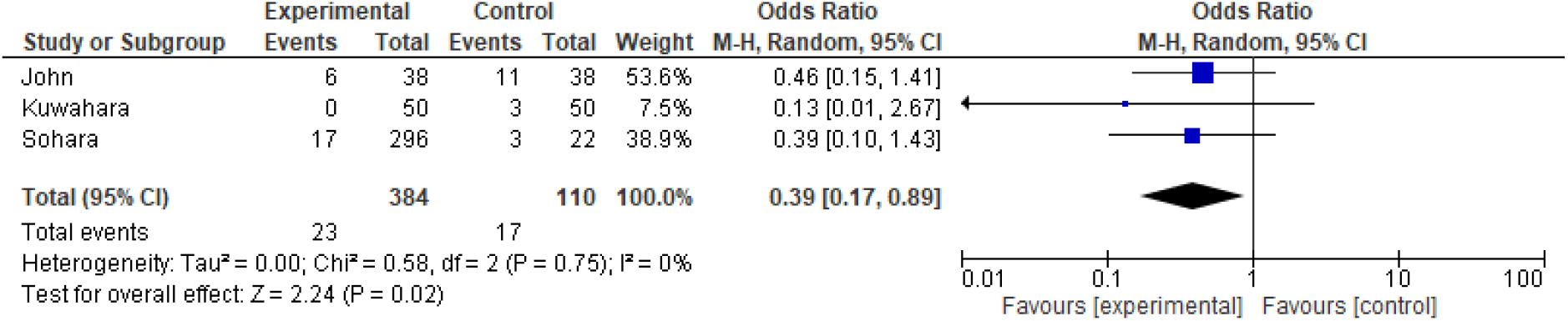
Forest plot of clinical studies comparing outcome of severe lesions. Events are the occurrence of grade III/IV lesions.

Finally, utilizing an ordinal logistic random intercept model rather than dichotomized outcome, we used GLIMMIX and NLMIXED in SAS to obtain an additional estimate of effect size using each category of lesion independently. This method showed that esophageal cooling by the method of direct instillation of cold water or saline used in these studies results in a point estimate of a -23% reduction in lesion grade, with 95% CI ranging from -85% to +38%.

## DISCUSSION

Esophageal injury from RF ablation remains a feared complication in the treatment of atrial fibrillation, and a variety of approaches have been developed to reduce this risk. Esophageal cooling has shown promise in a number of studies, generally utilizing low-capacity cooling approaches, such as direct instillation of cold water or saline directly into the esophagus via orogastric tube. Even with the limited heat extraction capacity of this approach, our meta-analysis of 3 studies suggests that cooling in this manner offers a clinically significant protective effect from severe lesions, and provides a 61% reduction in high grade lesion formation (with a 95% CI of 11% to 83% reduction).

Cooling via direct instillation of cold liquid into the esophagus is limited in actual heat extraction capacity by low volumes of direct liquid instillation. Even with the relatively high heat capacity of water, the total volumes (5mL to 20 mL at a time) serve as a significant limitation to the amount of thermal energy that can be absorbed. Higher heat extraction capacity has been evaluated in the past with various balloon device prototypes, where flow rates of 25 mL to 300 mL per minute were utilized. [13-16] These may offer greater heat extraction, and pre-clinical as well as mathematical models of early prototypes suggested benefits in lesion reduction, but none of these prototypes evolved into commercially-available products.

A device currently being investigated in clinical studies, designed for whole-body temperature manipulation, has a flow rate of up to 1900 mL per minute, far greater than any devices previously evaluated. This device is a commercially available esophageal cooling device utilized for general temperature management (inducing hypothermia, maintaining normothermia, or patient warming), and has shown protective effects in animal and mathematical models, as well as in recently presented clinical data.[21, 22]

A recent study on the prevalence and prevention of esophageal injury during atrial fibrillation ablation performed a meta-analysis of two studies of direct instillation of ice-cold water, but failed to distinguish between lesion severity and lesion frequency, focusing on the outcome of overall lesion frequency.[23] In contrast, our study had the benefit of an additional publication, and we analyzed data stratified by severity of lesions. Although a detailed understanding of the mechanisms of AEF formation is still developing, there is general agreement that thermal injury is a precursor, and that higher-grade thermal injury has higher risk of progression to AEF.[24]

Taking into account each lesion grade independently in a statistical model allows an alternative approach to estimate effect size that may provide further refinement of the estimate, at the cost of decreased precision. We found a point estimate using this approach suggesting a reduction in high-grade lesions of - 23%, although a higher number of patients with a greater number of high-grade lesions are necessary to narrow the confidence intervals around this estimate, which with the population included here, ranges from -85% to +38%. On the other hand, utilizing a methodology with greater performance and higher heat extraction capacity may increase the effect size point estimate further when either binary logistic or ordinal regression is utilized.

## LIMITATIONS

The studies analyzed in this report differed in specific RF techniques and equipment. Nevertheless, all used radiofrequency ablation, and the variation in technology reflects real-world practice currently. The studies reviewed did not all utilize randomization, and there was no description of any attempt at blinding the patients to the protection strategy used. Lesion grading varied between studies, but each scale involved a lesion characterization that allowed stratifying on common ground. Patient characteristics and ablation technologies and techniques utilized in the evaluated studies differed in many aspects; however, this may serve to broaden the generalizability of these findings.

## CONCLUSIONS

Esophageal cooling reduces lesion severity encountered during RF ablation, even when using relatively low heat extraction methods such as direct instillation of small amounts of cold liquid. Further investigation of this approach is warranted, particularly utilizing higher heat extraction technology such as closed-circuit devices with high coolant flow rates.

## Data Availability

Data are available publicly and upon request.

